# Risk assessment in cardiac surgery: Exploring machine learning and laboratory indices as adjunctive tools

**DOI:** 10.1101/2025.10.10.25337755

**Authors:** Wesley Chorney, John Hinchion

**Affiliations:** School of Medicine, University College Cork, County Cork, Ireland; Department of Cardiothoracic Surgery, Cork University Hospital, County Cork, Ireland

## Abstract

**Background:** Post-operative outcomes of cardiovascular surgery vary greatly among patients for a variety of reasons. While the specific reasons are often multifactorial and complex, certain machine learning methods are promising ways to both estimate mortality after surgery and elucidate important factors linked with mortality.

**Methods:** Using the MIMIC-IV database, we identified 11,261 patients on the cardiovascular surgery unit. We estimated all-cause one-year mortality for each patient after their most recent operation. The dataset included patient demographics (including age, sex, and BMI), as well as the maximum and minimum common laboratory values measured prior to each surgery (including electrolytes, eGFR, and red cell distribution width).

**Results:** Of the models tested, logistic regression outperformed all other approaches with respect to accuracy (*p* = 0.0075 with respect to a two-tailed *t*-test with the next strongest model). The model had an accuracy, sensitivity, and specificity of 85.07%, 82.89%, 85.19% respectively. Furthermore, features weighted heavily by the model are consistent with known predictors of mortality in the literature.

**Conclusion:** Pre-operative laboratory values are effective predictors of all-cause one-year mortality post-cardiovascular surgery used in conjunction with machine learning. Renal function, red cell distribution width, leukocytosis, and erythrocyte indices appear to be important prognostic factors.

## Introduction

The post-operative outcomes of cardiovascular surgery vary greatly, ranging from full recoveries to debilitating complications and death. Accurate understanding of the risks and benefits of surgery is therefore useful to both patients and physicians — for patients, this facilitates informed consent; while for physicians, this may aid in the decision to recommend surgery, or in planning post-operative care. To this end, tools to estimate the risk of surgery have been developed, with the European EuroSCORE II [1] and the American Society of Thoracic Surgeons (STS) score [2] being among the most widely used and well-studied.

Despite their ubiquity, these tools are not without limitations. For instance, the EuroSCORE II has been demonstrated to overestimate mortality in octogenarians [3]. More generally, the EuroSCORE II and the STS score tend to overestimate risk in high-risk populations, and underestimate risk in low-risk populations, respectively [4]. Possible reasons for these limitations include the type of data used in computing the score. While both scores include a number of patient factors and the type of operation, additional information could lead to stronger predictions.

Machine learning methods and statistical models offer alternatives to simpler scores. While complex machine learning methods often suffer from lacking explainability, certain models, such as random forests, are able to quantify how important a feature is in influencing the final prediction. Furthermore, other methods, such as Shapley values, can be used in conjunction with more complex models to increase explainability [5]. Complex architecture can also better deal with complex input. Consequently, these methods have been applied broadly in healthcare, such as in ECGs [6, 7], in X-ray interpretation [8, 9], and even as diagnostic aides for COVID-19 [10]. However, there has been a relative paucity of methods exploring and comparing machine learning methods in surgical fields. Models to predict mortality after cardiac surgery have been developed using pre-operative chest X-rays [11], the variables used to generate the EuroSCORE II (in which case the machine learning model showed a modest improvement) [12], both pre- and intra-operative variables [13], and pre-operative variables only [14–19]. Many of these models require large amounts of data — in some instances, independent variables are decided upon using feature selection from a larger set of hundreds of variables [17, 18]. In order to adapt these methods to new datasets with different features, such a process would have to be repeated.

In contrast to other methods, we show that patient demographics and common laboratory values can act as strong predictors of one-year mortality in patients undergoing cardiovascular surgery. In summary, we

- develop models to predict one-year mortality post-cardiovascular surgery using simple demographic information as well as common laboratory values,
- compare different machine learning models, and
- determine important predictors of mortality post-cardiovascular surgery.

## Materials and methods

### Dataset

Data used in this manuscript are from the MIMIC-IV dataset [20, 21], available freely from PhysioNet [22]. The dataset was accessed on 2025-04-12, and no information that could identify individual participants was accessed prior to, during, or after this date. The MIMIC-IV dataset contains data on 364,627 patients who either visited the emergency room or were admitted to the intensive care unit (ICU) at Beth Israel Deaconess Medical Center in Boston. Overall, these individuals had 546,028 hospitalizations and 94,458 ICU stays between 2008 and 2022. Patients are selected for inclusion if they are on the cardiovascular surgery service and undergo an ICD-9 or ICD-10 coded procedure related to the cardiovascular system during that time. For these patients, their demographic information, including age, sex, and BMI, basic medical information such as blood pressure and estimated glomerular filtration rate (eGFR), as well as the fifteen most frequently measured laboratory values across all patients in the dataset, are taken. These data are selected before the date of their respective operation, and for each laboratory value, both the minimum and maximum measured value are included in the dataset. Table 1 gives an overview of the patient demographics and basic medical information in the dataset. Altogether, the dataset is comprised of 37 features, including age, sex, number of previous operations, BMI, systolic blood pressure, diastolic blood pressure, eGFR, and the minimum and maximum values of bicarbonate, chloride, creatinine, hematocrit, hemoglobin, MCH, MCHC, MCV, platelet count, potassium, RDW, red blood cell count, sodium, urea nitrogen, and white blood cell count.

**Table 1.** Features of the dataset, reported as means with standard deviations. Only patients who had a recorded value for each attribute were included in the calculation. All values are reported as means with standard deviations in brackets.

| Attribute | Female ($n = 3296$) | Male ($n = 7965$) | Total ($n = 11261$) |
| --- | --- | --- | --- |
| Age | 67.55 (12.64) | 65.30 (11.58) | 65.96 (11.95) |
| BMI | 28.87 (7.31) | 28.77 (5.58) | 28.80 (6.20) |
| Systolic Blood Pressure | 133.28 (19.94) | 132.61 (18.91) | 132.82 (19.24) |
| Diastolic Blood Pressure | 73.01 (12.82) | 75.39 (12.55) | 74.65 (12.68) |
| eGFR | 41.63 (22.14) | 50.56 (20.97) | 46.80 (21.34) |
| Mortality | 6.64% | 4.29% | 4.98% |

Appendix A describes the most common operations (represented by ICD 9 and 10 codes) in the dataset, as well as how often they occur. We note that because patients were admitted to cardiac surgery, the operations they underwent were covered by multiple ICD codes — for instance, the most common item in Appendix A is extracorporeal circulation auxiliary to open heart surgery, but this would not be the only ICD code involved in the patient’s operation. The dataset consists of 804 unique procedures, determined by ICD 9 codes beginning with 35–38, and ICD 10 codes beginning with 02–06. Fig 1 gives a Kaplan-Meier survival curve for patients in the data. We note that the *y*-axis does not begin at zero for easier visibility. Overall, the mortality rate was 4.98%.

**Fig 1.**
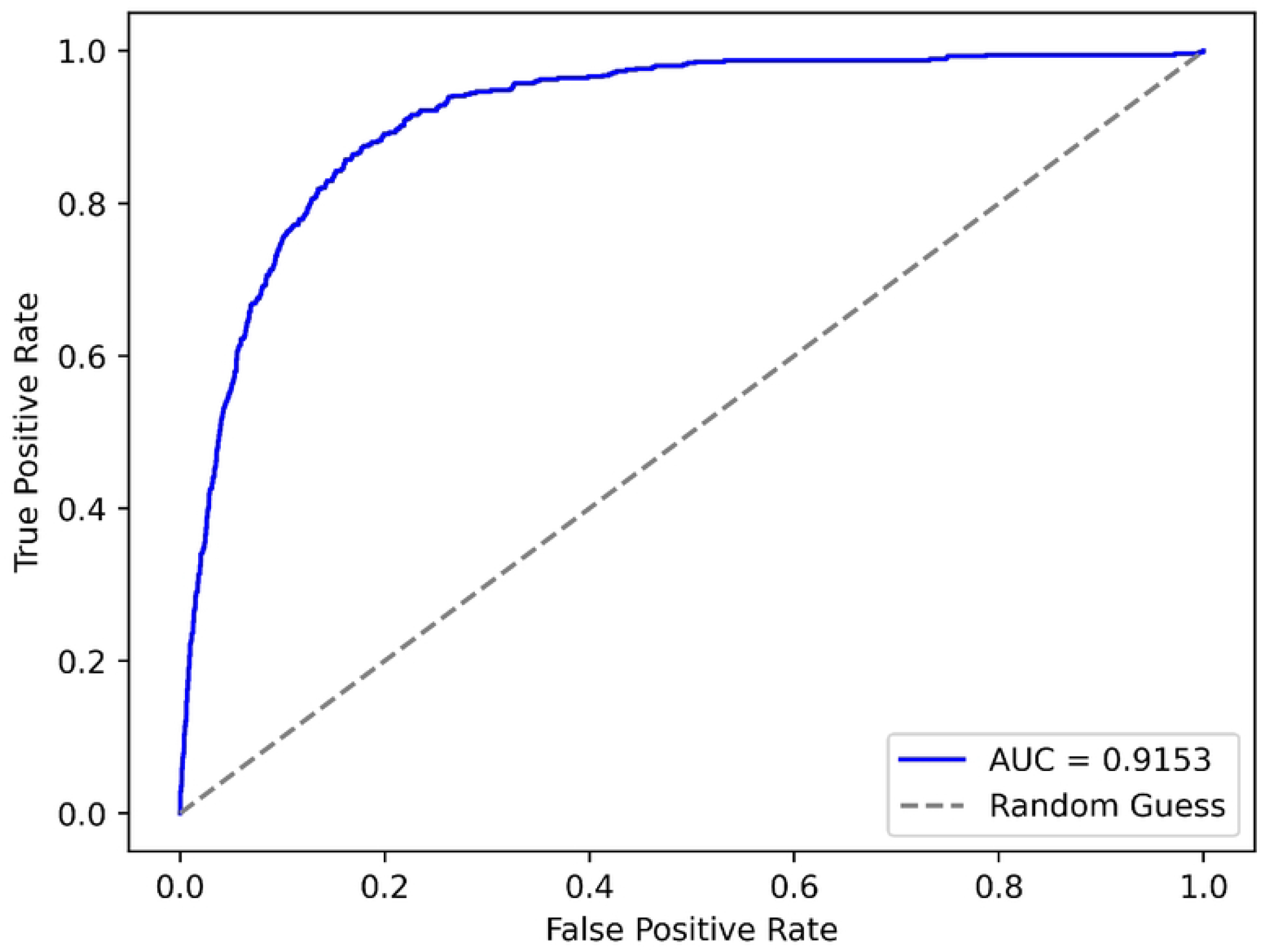
Kaplan-Meier survival curve for patients in the dataset. Note that for visibility, the *y*-axis does not begin at zero.

### Models

We test a variety of models, including random forest classifiers, extreme gradient boosted machines (XGBoost), support vector machines, neural networks, and logistic regression. Random forest classifiers are an ensemble method that combine a number of weak classifiers trained on a subset of the training data to create a stronger classifier. Ensembling allows for the reduction of bias while still allowing the model to adequately capture variance in the data [23]. Similar to random forest classifiers, extreme gradient boosted machines also function by combining weak predictors; however, in the case of gradient boosting, the predictors added at each iteration of the training address the residual error not captured by the existing predictors [24]. Support vector machines maximize the class-wise separation achieved by a hyperplane, and often make use of the so-called “kernel-trick” [25], where data is embedded in a higher dimension so that it can be more easily separated by a hyperplane. Finally, neural networks are nearly ubiquitous in machine learning for healthcare, and applications range from cardiology [26] to imaging [27] to evaluation of surgical skills [28]. We use a five-layer dense neural network, which is a sequence of matrices with nonlinear activation functions between each layer.

### Metrics

Typically, machine learning methods perform best on balanced data, though there are methods to address data imbalances. The dataset used in this work is heavily imbalanced, with one-year mortality of about 4.98% across all patients. In order to accurately assess each model, accuracy, sensitivity, and specificity are reported. Letting *TP, TN, FP, FN* denote true positives, true negatives, false positives, and false negatives, respectively, then

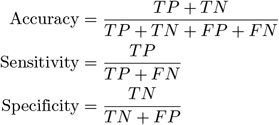

### Ethics

#### Human Participants

This study used de-identified data from the publicly available Medical Information Mart for Intensive Care IV (MIMIC-IV) database, hosted on PhysioNet [22]. The collection of patient information and creation of the MIMIC-IV resource were reviewed and approved by the Institutional Review Boards (IRBs) of the Massachusetts Institute of Technology (MIT, Cambridge, MA, USA) and Beth Israel Deaconess Medical Center (BIDMC, Boston, MA, USA) (MIT IRB #0403000206; BIDMC IRB #2001-P-001699/14), who granted a waiver of informed consent and authorized data sharing.

All patient data are fully de-identified in accordance with the Health Insurance Portability and Accountability Act (HIPAA) Privacy Rule. Consequently, the use of MIMIC-IV data for research purposes is considered exempt from additional institutional review board approval or informed consent requirements.

The required training to access the database was completed by the first author.

#### Participant Consent

This study did not involve direct interaction with human participants. The analysis was performed using the publicly available, de-identified Medical Information Mart for Intensive Care IV (MIMIC-IV) database.

The collection of patient data and creation of the MIMIC-IV resource were reviewed and approved by the Institutional Review Boards (IRBs) of the Massachusetts Institute of Technology (MIT, Cambridge, MA, USA) and Beth Israel Deaconess Medical Center (BIDMC, Boston, MA, USA) (MIT IRB #0403000206; BIDMC IRB #2001-P-001699/14). The IRBs granted a waiver of informed consent due to the retrospective nature of the study and the use of fully de-identified health records in compliance with the Health Insurance Portability and Accountability Act (HIPAA) Privacy Rule.

As the present study used only de-identified data, no additional participant consent was required.

## Results

Each model is trained and tested using stratified 5-fold cross validation. In the training phase, data was downsampled in order to achieve a more balanced training dataset, and was tested on the imbalanced test fold. Table 2 shows the results of each model across the five folds, presented as means plus or minus standard deviation.

**Table 2.** Overview of the results for each tested model. Values are reported as means plus or minus standard deviation, calculated across 5-fold stratified cross validation.

| Model | Accuracy | Sensitivity | Specificity |
| --- | --- | --- | --- |
| Logistic Regression | $85.07 \pm 1.11$ | $82.89 \pm 3.66$ | $85.19 \pm 1.03$ |
| Random Forest | $82.34 \pm 1.08$ | $87.34 \pm 1.97$ | $82.07 \pm 1.06$ |
| Support Vector Machine | $79.58 \pm 1.61$ | $80.57 \pm 3.42$ | $79.53 \pm 1.53$ |
| XGBoost | $82.87 \pm 0.32$ | $86.99 \pm 1.94$ | $82.65 \pm 0.30$ |
| Neural Network | $82.13 \pm 7.51$ | $77.69 \pm 15.49$ | $82.36 \pm 8.59$ |

**Table 3.** The most common procedures performed in patients in the dataset under consideration. We note that multiple procedure codes may be listed per operation.

| Operation | Count |
| --- | --- |
| Extracorporeal circulation auxiliary to open heart surgery | 3521 |
| Bypass Coronary Artery, One Artery from Left Internal Mammary, Open Approach | 2668 |
| Single internal mammary-coronary artery bypass | 2157 |
| Excision of Left Saphenous Vein, Percutaneous Endoscopic Approach | 1502 |
| Insertion of Infusion Device into Superior Vena Cava, Percutaneous Approach | 952 |
| Bypass Coronary Artery, Two Arteries from Aorta with Autologous Venous Tissue, Open Approach | 930 |
| Replacement of Aortic Valve with Zooplastic Tissue, Open Approach | 892 |
| Excision of Right Saphenous Vein, Percutaneous Endoscopic Approach | 866 |
| Open and other replacement of aortic valve with tissue graft | 857 |
| (Aorto)coronary bypass of two coronary arteries | 788 |
| (Aorto)coronary bypass of three coronary arteries | 750 |
| Venous catheterization, not elsewhere classified | 725 |
| Bypass Coronary Artery, One Artery from Aorta with Autologous Venous Tissue, Open Approach | 641 |
| Bypass Coronary Artery, Three Arteries from Aorta with Autologous Venous Tissue, Open Approach | 628 |
| Excision of Left Radial Artery, Percutaneous Endoscopic Approach | 553 |
| Bypass Coronary Artery, One Artery from Aorta with Autologous Arterial Tissue, Open Approach | 503 |
| (Aorto)coronary bypass of one coronary artery | 474 |
| Supplement Mitral Valve with Synthetic Substitute, Open Approach | 451 |
| Bypass Coronary Artery, One Artery from Right Internal Mammary, Open Approach | 407 |
| Central venous catheter placement with guidance | 329 |
| Bypass Coronary Artery, Two Arteries from Left Internal Mammary, Open Approach | 324 |
| Excision of Mitral Valve, Open Approach | 298 |
| Replacement of Aortic Valve with Synthetic Substitute, Open Approach | 284 |
| Open heart valvuloplasty of mitral valve without replacement | 284 |
| Replacement of Thoracic Aorta, Ascending/Arch with Synthetic Substitute, Open Approach | 262 |
| Destruction of Conduction Mechanism, Open Approach | 262 |
| Occlusion of Left Atrial Appendage with Extraluminal Device, Open Approach | 261 |
| Open and other replacement of aortic valve | 256 |
| (Aorto)coronary bypass of four or more coronary arteries | 243 |
| Resection of vessel with replacement, thoracic vessels | 233 |
| Left heart cardiac catheterization | 196 |
| Insertion of Infusion Device into Right Atrium, Percutaneous Approach | 177 |
| Hemodialysis | 172 |
| Replacement of Mitral Valve with Zooplastic Tissue, Open Approach | 167 |
| Insertion of Monitoring Device into Upper Artery, Percutaneous Approach | 164 |
| Insertion of Pacemaker Lead into Right Ventricle, Percutaneous Approach | 159 |
| Bypass Coronary Artery, Four or More Arteries from Aorta with Autologous Venous Tissue, Open Approach | 152 |
| Arterial catheterization | 145 |
| Insertion of Pacemaker Lead into Right Atrium, Percutaneous Approach | 145 |
| Supplement Tricuspid Valve with Synthetic Substitute, Open Approach | 145 |
| Excision, destruction, or exclusion of left atrial appendage (LAA) | 139 |
| Cardioplegia | 126 |
| Excision or destruction of other lesion or tissue of heart, open approach | 126 |
| Insertion of drug-eluting coronary artery stent(s) | 126 |
| Annuloplasty | 119 |
| Intraoperative cardiac pacemaker | 118 |
| Open and other replacement of mitral valve with tissue graft | 110 |
| Initial insertion of dual-chamber device | 104 |
| Initial insertion of transvenous leads [electrodes] into atrium and ventricle | 101 |
| Venous catheterization for renal dialysis | 98 |

We note in particular that the logistic regression classifier is the best performing model with respect to accuracy, and is well-balanced with respect to sensitivity and specificity. For this reason, we investigate the logistic regression classifier closer, including its coefficients (which relate to the relative importance of a feature in predicting the null or positive class).

Fig 2 is the receiver-operating characteristic (ROC) curve (as well as the area underneath the curve) for the logistic regression classifier. In addition, the receiver-operating characteristic curve for a random guess classifier (which would achieve an area under the ROC curve of 0.5) is shown as a dashed line. The curve demonstrates how the true and false positive rate vary as the threshold for positive class estimation is varied. A higher area under the ROC curve represents a stronger classifier.

**Fig 2.**
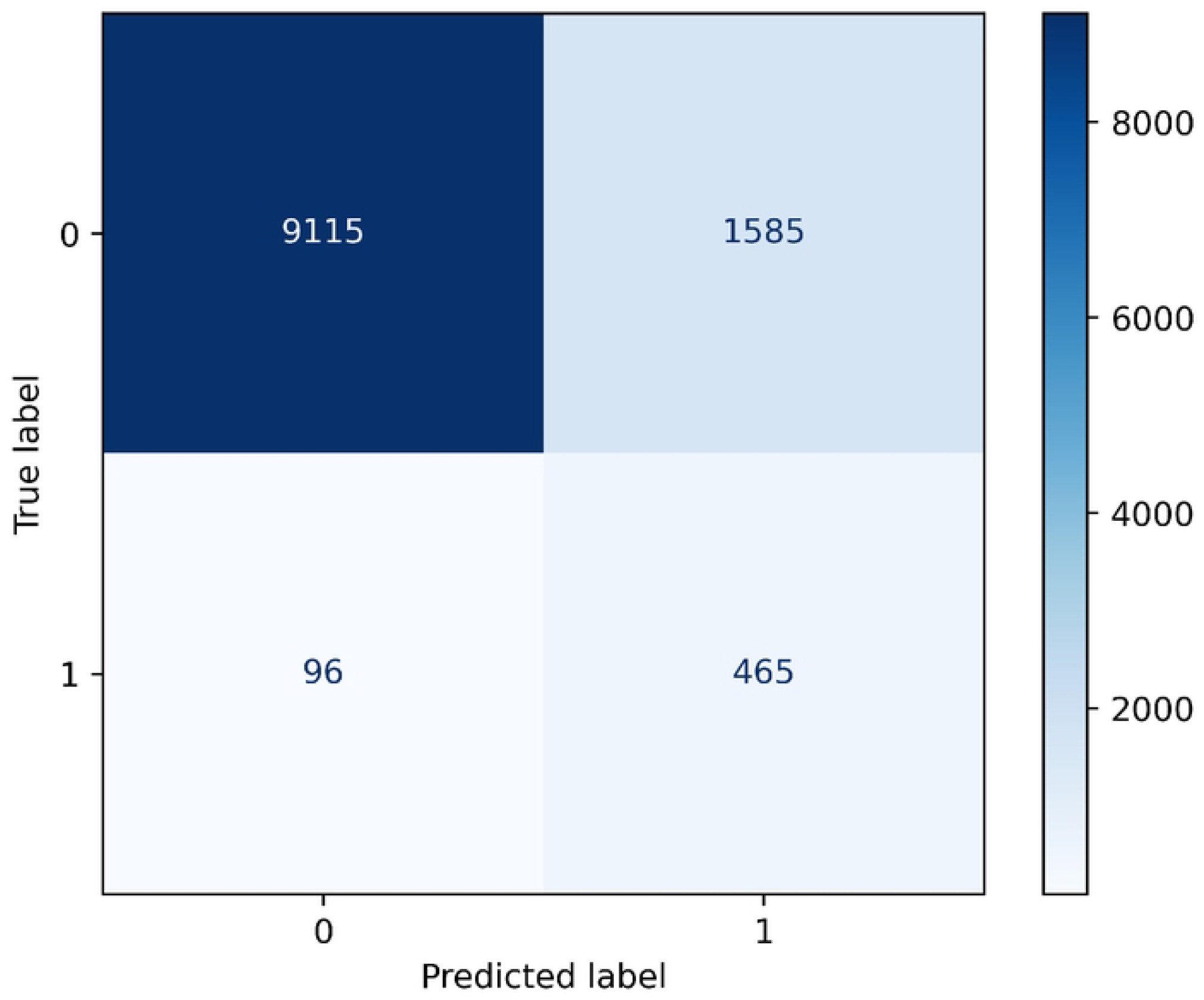
Area under the receiver-operating characteristic curve for the logistic regression classifier. Another receiver-operating characteristic curve is plotted in a dashed line for a random guess classifier for comparison.

Fig 3 displays the confusion matrix for the logistic regression classifier. Note in particular the imbalanced nature of the dataset, with one-year all-cause mortality present in approximately 4.98% of patients. Fig 4 presents the predicted probability distribution for the null (negative) class and the positive (one-year mortality) class. In general, the model is relatively discriminative, but including additional features in the model could improve its ability to discriminate between positive and negative classes.

**Fig 3.**
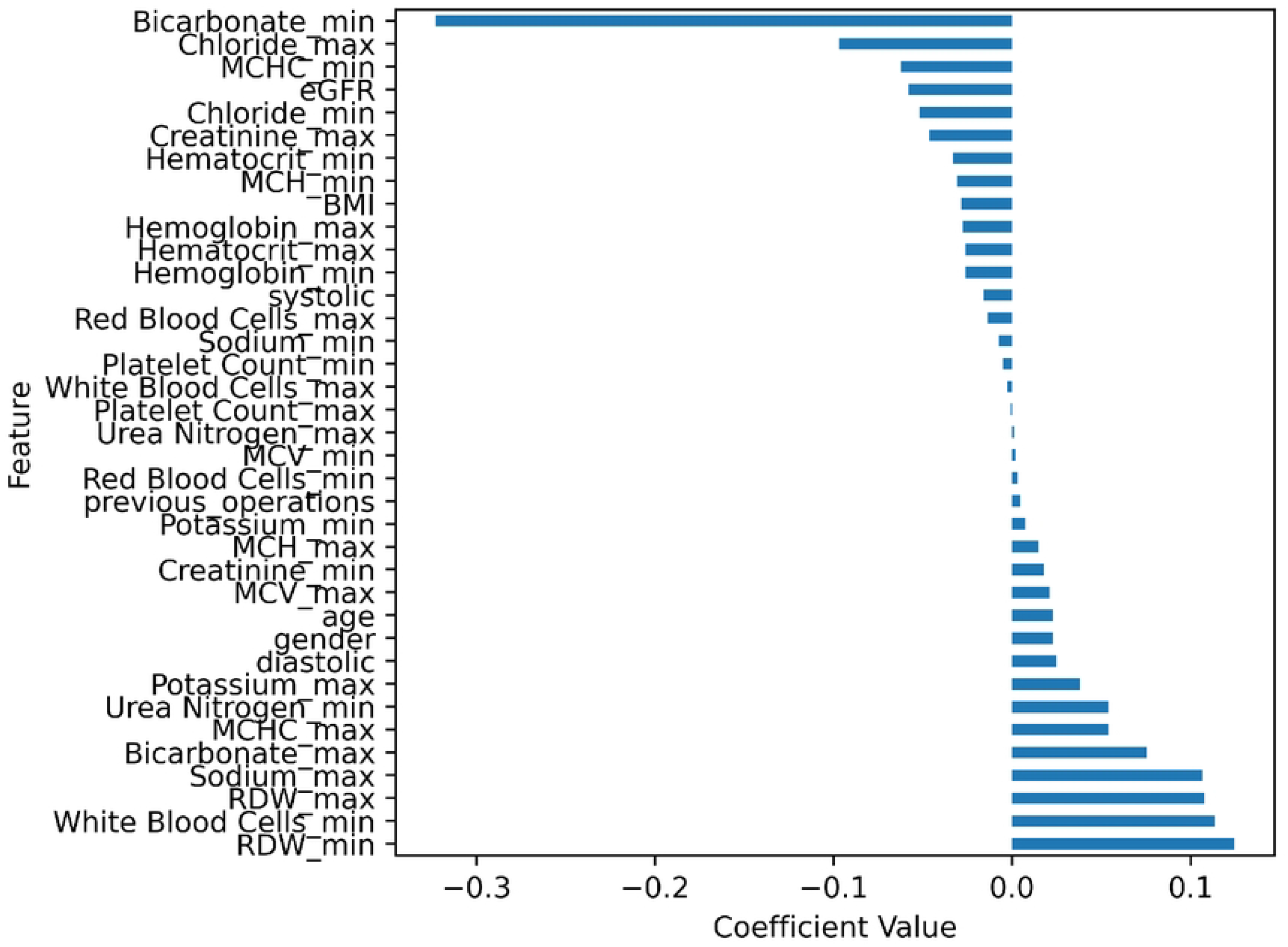
Confusion matrix for the logistic regression classifier.

**Fig 4.**
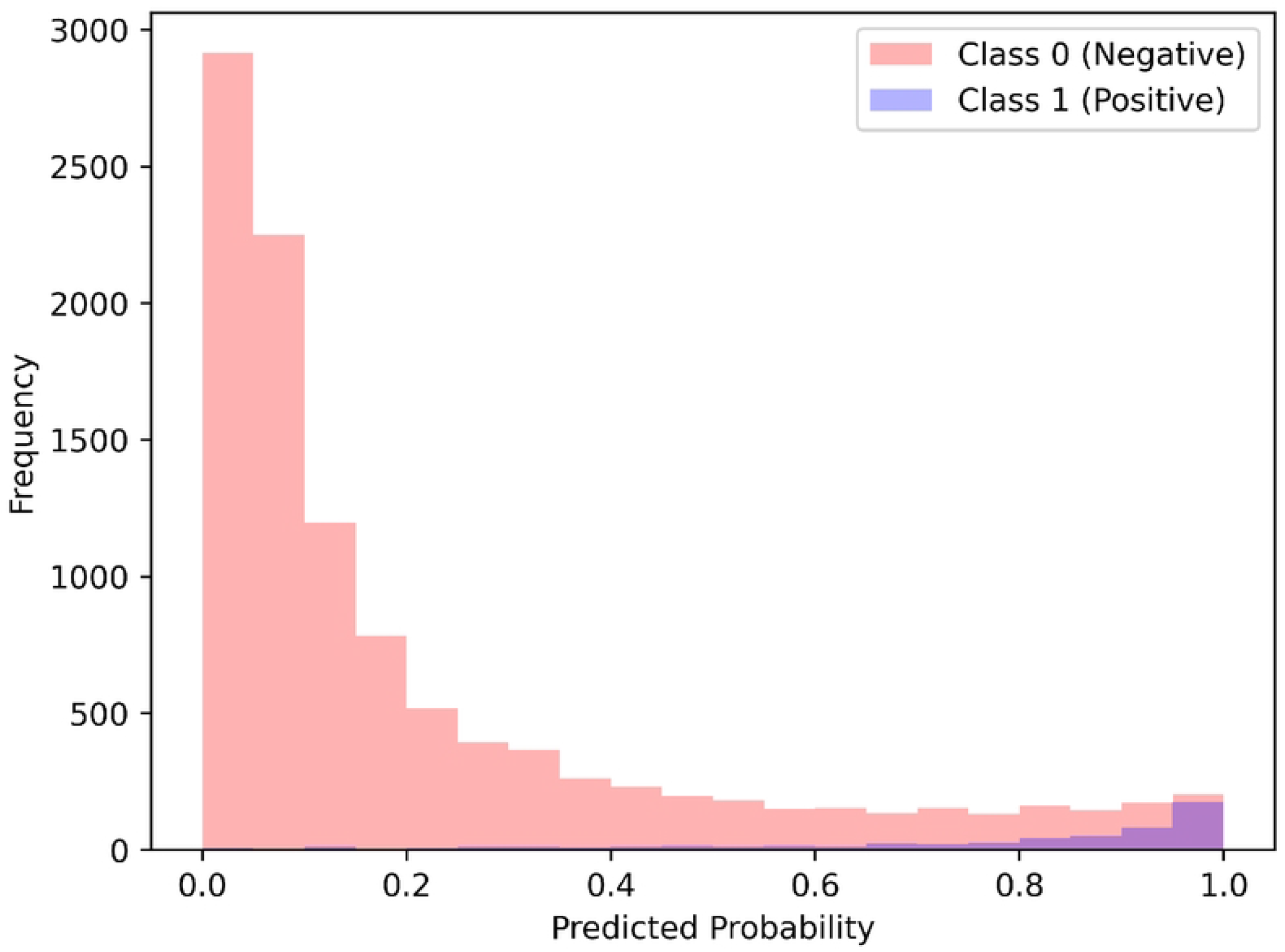
Distribution of predicted probabilities for both the null class (survivors after one year) and the positive class (patients who died up to one year post-cardiovascular surgery).

Finally, Fig 5 displays the coefficients for each feature in the logistic regression model. Given that all features are positive, the coefficient values indicate the contribution of each feature to the prediction of the null or positive class. Recall that the coefficients *c*_*i*_ and features *f*_*i*_ of a model define a value 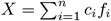, which is then used in a sigmoid function to generate a probability 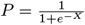. Therefore, the coefficients of a logistic regression model do not have as convenient an interpretation as in linear regression, but a unit increase in *f*_*i*_ does increase *X* by *c*_*i*_ (assuming all other features are kept constant). Practically, higher values for a negative coefficient lead to a higher predicted probability of surviving after cardiac surgery (for instance, having a higher minimum bicarbonate value appears to be a strong predictor for survival), whereas higher values for positive coefficients lead to a lower predicted probability of survival.

**Fig 5.**
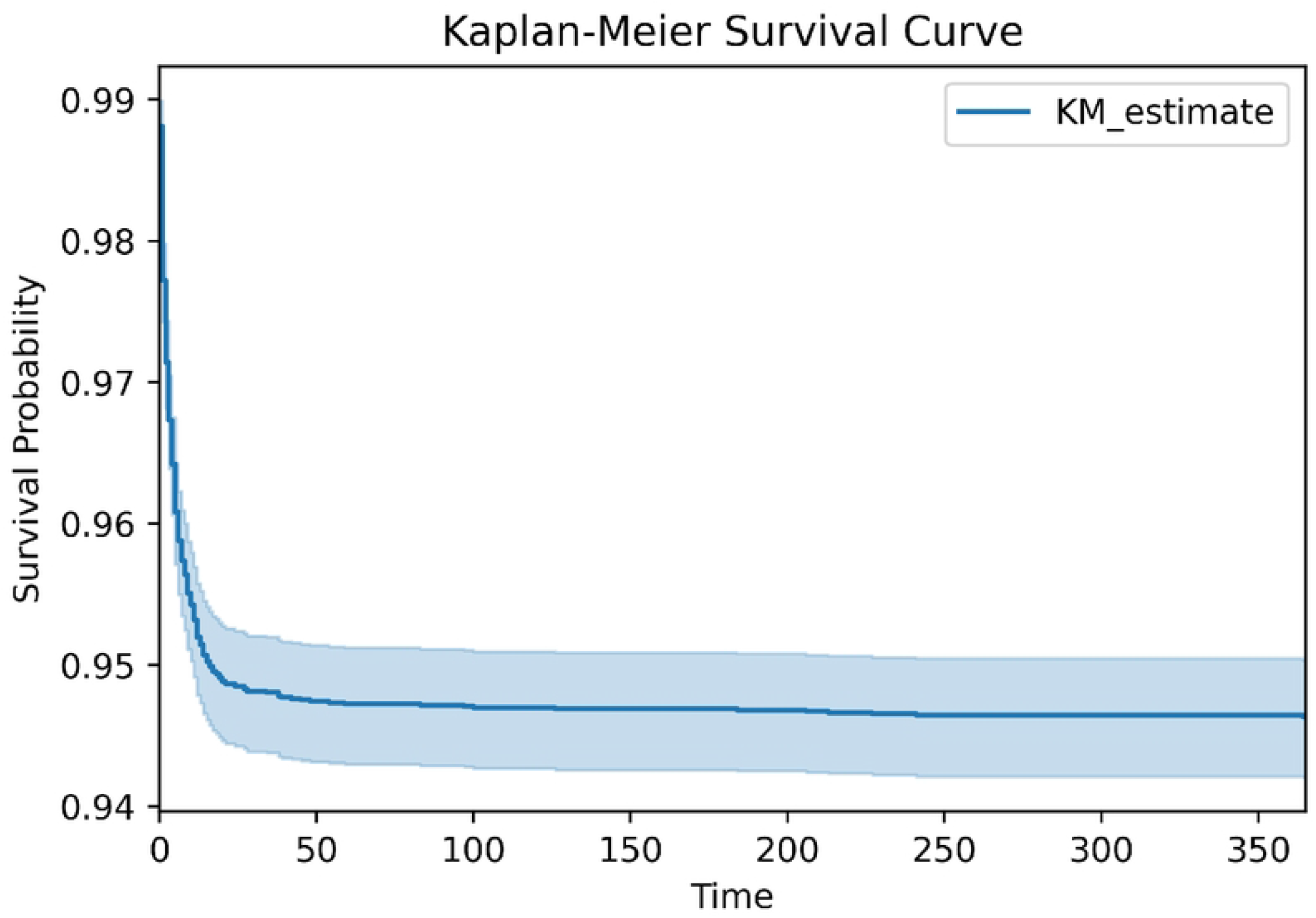
Coefficients for the linear regression classifier. The value of a coefficient correlates with its contribution to the prediction of survival or perishing.

## Stroke Subgroup Analysis

Given that stroke is a significant postoperative complication [29], we investigate how the proposed model performs on those patients who develop a stroke within one year of their operation. A stroke is detected in the dataset as ICD 9 codes of 430–434 or 436–438 (transient ischemic attacks are excluded), or ICD 10 codes with the roots I60–I64. In total, there are 66 patients with a diagnosis of stroke within a year after their operation. On these 66 patients, the model performs with an accuracy, sensitivity, and specificity of 65.2%, 75.0%, 63.8%, respectively.

We note that this subgroup analysis is limited for a number of reasons. First, the stroke rate is approximately 0.5%, which falls outside of the range typically quoted in literature [30]. This is most likely due to data in MIMIC-IV capturing only admissions to a single medical center; whereas patients who had their operation at Beth Israel Deaconess Medical Center and were subsequently admitted to a different center for stroke care would not be included in the subgroup of patients with stroke. Therefore, the subgroup is not an accurate representation of incidence of stroke post-cardiac surgery. Furthermore, more patients would be needed to provide a more accurate estimate of model performance. Second, while stroke is a significant complication with high mortality [31], it is distinct from mortality, and hence represents a cohort of patients sampled from a different distribution than expected by the model. However, future work could investigate both the prediction of stroke post-cardiovascular surgery, as well as mortality prediction in those patients who develop stroke post-cardiovascular surgery.

## Discussion

RDW and poor post-operative outcomes, particularly mortality, are well-known to be correlated in both non-cardiac [32, 33] and cardiac [34] surgery. This is well reflected in the model, as both the minimum measured and maximum measured pre-operative RDW have large positive coefficients. Similarly, leukocytosis is a well-known marker of non-specific inflammation, and has been shown to be related to adverse outcomes post-cardiac surgery [35]. The logistic regression model also heavily favours indicators of good kidney function — minimum pre-operative bicarbonate is associated with a strongly negative coefficient, as well as eGFR (to a lesser extent). Given that cardiac surgery is associated with acute kidney injury [36], it is likely that normal renal function improves mortality. For instance, Thakar et al. demonstrated that pre-operative renal function, as measured by GFR, attenuates the effect of post-operative renal failure on mortality [37]. Finally, we also find that erythrocyte indices are partitioned by the model — a higher minimum pre-operative MCV, MCHC, and MCH, as well as a lower maximum pre-operative MCV, MCHC, and MCH, are associated with better outcomes post-cardiac surgery. Given the well-known relation between anemia and post-operative mortality in cardiac surgery [38], this is unsurprising. Overall, we find that the model agrees well with the literature.

It is known that combining (or *ensembling*) classifiers can reduce bias in a model [23]. Typically, these classifiers are trained on different features or with different objectives, as in the case of random forest or gradient boosted models. Similarly, the proposed model could be used in conjunction with the STS score and the EuroSCORE II, or with other mortality prediction models, in order to improve the overall estimate of risk of mortality. Both the STS score and the EuroSCORE II focus on indicators of cardiovascular health, demographics, surgery type, and other risk factors such as pulmonary function, and thus the proposed model is likely sufficiently different to complement these scores. However, further work would be needed to establish this. In addition, a more detailed dataset would be required so that the STS score and the EuroSCORE II could also be computed for each patient.

While the purpose of the current study was to evaluate how common pre-operative laboratory values could be used to estimate one-year mortality post-cardiovascular surgery, the proposed model could be expanded to include additional factors. For example, different operations carry with them different risks, and thus the specific type of operation being performed plausibly contains very important prognostic information. Other features that are included in the STS score or the EuroSCORE II would most likely benefit the model as well, such as cardiovascular or pulmonary risk factors. However, if these features were to be included in the model, then it would most likely be less useful to combine the resultant model with the STS score or the EuroSCORE II, since the overlap in inputs would be large, and both rely on logistic regression.

It is of note that logistic regression does not model interactions between features, while other models, such as neural networks, are able to do so. Nevertheless, the logistic regression classifier performed the best of the tested models. There are various possible explanations for this. First, it could be the case that there are no significant interactions between variables. For instance, while chloride is an osmotically active element [39], and thus could affect systolic blood pressure, the effect of minimum chloride on systolic blood pressure may be attenuated by other means of balancing osmolarity. In this case, the interaction between the two features could be negligible, and thus provide little additional information to a model. Furthermore, the given dataset is much smaller than most datasets used in conjunction with neural networks and other complex models. Therefore, more complex patterns between variables may not be adequately modeled. For instance, note that the number of *m*th order interactions between *n* variables scales by *n*^*m*^, which quickly expands past the size of the dataset. Perhaps most likely is some combination of the two explanations leading to logistic regression being the strongest classifier.

Given that the study focused on commonly recorded lab values, the approach could be easily adopted in many settings. Furthermore, given that the classifier is relatively simple, the approach could easily be deployed in resource constrained settings. However, the given model was trained on patients from the MIMIC-IV dataset who were admitted to the cardiac surgery unit, and the MIMIC-IV dataset consists of patients initially admitted to the intensive care unit or emergency department at Beth Israel Deaconess Medical Center. Therefore, the generalizability of the current study may be limited by the patient population, inasmuch as diseases vary by geographic location [40], and the type of presentation, since these patients were either admitted to the intensive care unit or emergency department. Therefore, further testing would be required to determine whether or not the model performance as reported would be accurate. Nevertheless, the fact that the model seems to prioritize indicators which are well-established in the literature is encouraging.

The clinical applicability of the proposed model is twofold. First, it is clear that better pre-operative mortality predictions could help in more accurately weighing risks and benefits for a procedure. Combining the proposed model with the STS score or the EuroSCORE II may help improve the mortality predictions from either score alone. The features required by the model are also extremely basic. A second benefit is that due to its simplicity, the model is explainable. The impact of a unit change in each feature can be seen by the magnitude of its corresponding coefficient, and while these impacts will be amplified or attenuated based on the other features, coefficients can be compared to better understand relative feature importance. These features could then be optimized prior to operation. However, though we believe that the proposed model could generate clinical benefit, we do not believe that it could replace the clinical judgment garnered over years of education and experience, but rather could complement this judgment.

## Conclusion

Pre-operative laboratory values are commonly recorded prior to surgery. However, whether they are being fully utilized with respect to gauging whether a patient is suitable for surgery is unclear. While the STS score and the EuroSCORE II do look at some of these values, we demonstrated that simple machine learning models could be used to generate a prediction of all-cause one-year mortality post-cardiovascular surgery. The simple nature of the proposed model allows for a high degree of explainability, and the coefficients of the model weigh heavily those features that are indicated as important by the literature.

Overall, we have shown that pre-operative laboratory values have potential to complement existing mortality scores to better predict mortality post-cardiovascular surgery. While the current study focused on a sizable cohort of patients from the MIMIC-IV dataset, further work could investigate whether our findings generalize to other patient populations. Larger datasets may also allow for more complex machine learning models capable of more accurate predictions.

## Data Availability

Data cannot be shared publicly because it is derived from the MIMIC-IV dataset, which requires training before access. Access to the dataset is granted via PhysioNet for researchers who successfully complete the training.

https://physionet.org/content/mimiciv/3.1/

## Appendix A

